# Navigating primary health care challenges: Insights from older people with multimorbidity in Malawi

**DOI:** 10.1101/2024.03.22.24304706

**Authors:** Duncan Kwaitana, Maya Jane Bates, Esnath Msowoya, Dorothee van Breevoort, Thomas Mildestvedt, Eivind Meland, Eric Umar

## Abstract

The global population is undergoing a significant surge in aging leading to increased susceptibility to various forms of progressive illnesses. This phenomenon significantly impacts both individual health and healthcare systems. Low and Middle Income Countries face particular challenges, as their Primary Health Care (PHC) settings often lack the necessary human and material resources to effectively address the escalating healthcare demands of the older people. This study set out to explore the experiences of older people living with progressive multimorbidity in accessing PHC services in Malawi. Between July 2022 and January 2023, a total of sixty in-depth interviews were conducted with dyads of individuals aged ≥50 years and their caregivers, and twelve healthcare workers in three public hospitals across Malawi’s three administrative regions. The study employed a stratified selection of sites, ensuring representation from rural, peri-urban, and urban settings, allowing for a comprehensive comparison of diverse perspectives. Guided by the Andersen-Newman theoretical framework, the study assessed the barriers, facilitators, and need factors influencing PHC service utilization by the older people. Three themes, consistent across all sites emerged, encompassing barriers, facilitators, and need factors respectively. The themes include: (1) clinic environment: inconvenient clinic setup, reliable PHC services and research in diabetic foods; (2) geographical factors: bad road conditions, lack of comprehensive PHC at local health facility and need for community approaches; and (3) social factors: encompassing use of alternative medicine, transport support, perceived health care benefit and support with startup capital for small-scale businesses. This research highlights the impact of multiple factors on the access to and utilization of PHC services among older individuals, emphasizing the urgent need for enhanced nationwide availability of such services. It strongly recommends a thorough investigation into successful practices implemented in diverse health facilities in Malawi, with a specific focus on addressing the unique healthcare needs of the older population.

## Introduction

In an era marked by unforeseeable public health hurdles, one undeniable trend emerges—the global population is aging swiftly [1,2]. Multimorbidity, which refers to the concurrent existence of multiple chronic conditions, becomes more prevalent as individuals age [3]. In the context of research in Africa, the World Health Organization (WHO) classifies older people as individuals aged ≥50 years [4].

Worldwide, numerous healthcare systems were originally structured to cater to a relatively youthful population, prioritizing curative care aligned with different health needs than those currently encountered by the older populations [5,6]. This has created numerous challenges for the older people to easily access Primary Health Care (PHC) services. Primarily, older people in Low and Middle Income Countries (LMICs), such as Malawi, often face challenges in accessing primary healthcare services. This difficulty arises, among others, from the lack of designated clinics specifically catering to their needs [7].

The presence of multimorbidity, compounded by advanced age, leads to significantly increased healthcare utilization and elevated social care costs [8]. Community health care facilities face significant challenges in delivering comprehensive PHC services across many LMICs [5,9–11]. These challenges stem from the inherent weaknesses in PHC systems, marked by severe shortages in healthcare infrastructure, human resources, as well as essential medicines and supplies [12]. A recent systematic review evaluating the access and utilization of PHC services for older individuals across LMICs identified various additional barriers [9]. These barriers encompassed negative healthcare attitudes, considerable distances to PHC facilities, extended waiting times, insufficient integration of PHC with other services, and instances of stigma and discrimination. Consequently, a significant proportion of older people experiencing progressive multimorbidity fail to either access or fully realize the optimum advantages of utilizing PHC services, whether at the community or higher-level facilities. This, in turn, adversely contributes to the deterioration of their health, and in the worst cases, may result in preventable deaths [13].

Malawi, classified as a low-income country in Southern Africa, is home to a population of approximately 17.6 million people [14]. There is paucity of evidence regarding experiences of older people living with multimorbidity in accessing PHC services in the Sub-Saharan Africa [15]. However, the United Nations (UN) emphasizes the necessity of aligning health and long-term care systems to effectively address the requirements of aging populations. [16]. This study, therefore, set out to explore the experiences of older individuals with multimorbidity, particularly their access to and utilization of PHC and expectations for improved service delivery in Malawi.

## Methods

### Study design

This is a PHC services research that employed a cross sectional qualitative phenomenological design. This approach was chosen in order to elucidate the core of a phenomenon by examining it through the lens of those who undergo it, with the aim of comprehending the significance that participants attribute to the phenomenon [17]. In addition, we used exploratory interviews to facilitate the assessment of the factors affecting access and utilization of PHC services by older people living with progressive multimorbidity. The study was conducted from 18 July 2022 to 02 January 2023.

### General setting: South, Centre and North regions of Malawi

Malawi is divided into 28 districts within three major administrative regions, namely: South, Center and North [18]. This study was carried out in three districts, each representing one of the three regions and featuring the presence of PHC. In the South, the study site was at Mangochi district hospital, a 500-bed secondary and public health facility. Mangochi is a peri-urban district with a projected 2023 population of 1,346,740 inhabitants [18]. In the Central region, the study site was at Kamuzu Central Hospital, a tertiary government referral facility with a bed capacity of 1,250 and serving approximately 5 million people annually [19]. Kamuzu Central Hospital is in Lilongwe, the capital city of Malawi. Mzimba district hospital (∼274-bed capacity), another secondary and public health facility, was the third study site located in a rural district of Mzimba in the Northern region of Malawi serving a total population of 940,184 persons [14]. The use of a stratified selection for study sites facilitated the creation of a representative sample for Malawi, taking into account different places of settlement: rural, peri-urban, and urban. This also allowed for a diverse comparison of experiences of participants in view of primary healthcare services influenced by cultural, social and spiritual factors.

Public healthcare services in Malawi are offered free of charge. These facilities primarily cater to the general population, and there are currently no public or private facilities specifically designated for the older people in the country [7]. There are only a few modest community initiatives aimed at older individuals, and these initiatives are predominantly associated with the activities of civil society or non-governmental organizations [7,20].

### Participant selection

A purposive sampling frame was used to assess experiences around access and utilization of primary healthcare services by the older people living with progressive multimorbidity. There was a total of 60 adult patients and caregivers (n = 20 at Mangochi District Hospital, n = 20 at Kamuzu Central Hospital, n = 20 at Mzimba District Hospital) who enrolled in this study and participated in in-depth interviews. Additionally, a total of 12 healthcare worker qualitative interviews (n = 4 at Mangochi District Hospital, n = 4 at Kamuzu Central Hospital, n = 4 at Mzimba District Hospital) were conducted across all the three sites. Information-rich cases were those with the ability to provide contextual experiences from which one could learn a great deal about issues of central importance in line with objectives of the research project. **Table 1** presents characteristics of study participants.

**Table 1.** Characteristics of older people/caregiver dyads and healthcare workers participating in qualitative interviews at Mangochi and Mzimba District Hospitals and Kamuzu Central Hospital, July 2022 - January 2023.

| No. | District/Region | Name of Health Facility | Category | Participant Characteristics | n = 72 (F = 39) |
| --- | --- | --- | --- | --- | --- |
| 1 | Mzimba - North | Mzimba District Hospital | Rural | Older people with multi-morbidity | 10 (6) |
|  |  |  |  | Carer givers of older people | 10 (8) |
|  |  |  |  | Healthcare service providers | 4 (3) |
| 2 | Mangochi - South | Mangochi District Hospital | Peri-urban | Older people with multi-morbidity | 10 (9) |
|  |  |  |  | Carer givers of older people | 10 (6) |
|  |  |  |  | Healthcare service providers | 4 (2) |
| 3 | Lilongwe - Centre | Kamuzu Central Hospital | Urban | Older people with multi-morbidity | 10 (7) |
|  |  |  |  | Carer givers of older people | 10 (8) |
|  |  |  |  | Healthcare service providers | 4 (0) |

F: Female

### Study Theoretical Framework

Our analysis draws on the Andersen-Newman theoretical framework of health services utilization (**S1. Fig 1**) [21]. The framework predicts that a series of factors; predisposing, enabling and need factors influence the utilization of health services by people. In this paper, we focus on the facilitating factors, barriers and need factors. Adopting the framework helped to identify a series of factors that influenced or impeded access and utilization of PHC services by older people living with progressive multimorbidity. Additionally, older people managed to express their needs to enhance access and utilization of PHC services.

### Data collection

Hospital management teams from the respective study sites were briefed about the study; its objectives and methodologies, and potential contribution of the findings to policy and practice. Well-trained Research Assistants (RAs) were deployed to conduct the recruitment processes. COVID-19 preventive measures were adhered to including; wearing of face masks, observing social and physical distance, regular hand sanitization or hand washing using soap. The RAs visited the study sites during clinic days for the various chronic conditions for pre-screening of potential participants with support from hospital focal point persons. Through the same process, separate arrangements were made where some participants were recruited from their respective communities. Participants that fulfilled the inclusion criteria were briefed about the study, and asked if they would like to participate. Those who expressed interest were requested to provide written consent before being enrolled into the study. Participants were informed that it was allowable for them to refuse to participate in the study or withdraw from the study at any time without facing any consequences.

Exploratory in-depth qualitative interviews were then carried out with the eligible participants (older people and caregiver dyads and healthcare workers) simultaneously to assess their experiences around primary healthcare access and utilization. The interviews were conducted in any of the following local languages; Chichewa, Yao or Tumbuka including English especially with healthcare workers. To ensure rigor in data collection and analysis, all the interviews were digital recorded.

### Data management

Data audio recordings were transcribed verbatim by research assistants. The interviews that were conducted in local languages with older people and caregiver dyads were translated during transcription. Interviews with health care workers were conducted in English, and therefore did not require translating. Unique codes were assigned to each participant for anonymity of data and enhanced ease of identification. All study related tools in hard copies were kept in lockable cabinets for purposes of data safety and only accessible by the researchers. Similarly, a password protected laptop was used to maximize safety of storage of all study data and only the principal investigator had the right of access. Two study team members (DK and EsMs) thoroughly reviewed all transcripts to ensure their completeness.

### Data analysis

The analysis began with data emersion, where two researchers (DK and EsMs), independently read the transcripts multiple times. This process was done inductively with the aim of comprehending the data and pinpoint emerging themes and sub-themes that captured the experiences of study participants in relation to PHC access and utilization. Subsequently, the two researchers convened multiple times to deliberate on the identified themes and sub-themes, reaching a consensus on the development of a code book. The formulation of the code book was guided by the deductive Andersen-Newman thematic framework analytical approach, which emphasizes barriers, facilitators, and need factors related to accessing healthcare services. The transcripts were then imported into NVIVO version 12 pro and systematically coded by DK. The results of the data analysis process were deliberated upon during a study team meeting with the rest of the researchers.

### Ethical considerations

Ethical approval for the study was granted by the College of Medicine Research and Ethics Committee (COMREC) with the assigned application number P.05/22/3646. All participants were provided with an information sheet and asked to provide written informed consent or using thumb print if required. A distress protocol (**S2**) was developed to enable participants to pause or terminate interviews wherever necessary. Recognizing that this interview would explore sensitive personal experiences with the potential to elicit emotional responses from participants, a distress protocol assists researchers in navigating these situations responsibly [22]. This ensures that participants are not unduly harmed or distressed and are provided with proper assistance as needed. Data collected for the study was anonymized using participant identification numbers. All person identifying information was kept separate from participant files.

## Results

A total of thirty older person/caregiver dyads (n = 60) were recruited from across the three study sites, which were all public health facilities. Individuals in the older age group ranged from 54 to 89 years, whereas caregivers spanned in age from 20 to 82 years. Twelve healthcare professionals, each with a minimum of six months of experience in PHC settings across the study sites, participated in this study.

Our findings unveiled three major themes: (I) clinic environment (II) geographical factors, (III) social factors. These themes functioned as key barriers, facilitators and need factors for accessing PHC services. Below, we elaborate on these thematic areas in the context of facilitators, barriers and need factors, and how they impacted older people in accessing PHC. The corresponding data is depicted in Table 2.

**Table 2.** Thematic Areas underpinned by Andersen-Newman Theoretical Framework.

| No. | Major themes | Facilitators | Barriers | Need factors |
| --- | --- | --- | --- | --- |
| 1 | Clinic environment | Reliable PHC services | Inconvenient clinic setup | Research on diabetic foods |
| 2 | Geographical factors |  | Bad road conditions | Community approaches |
|  |  |  | Lack of comprehensive PHC at the local facility |  |
| 3 | Social factors | Available transportation support | Use of alternative medicine | Support with startup capital for business |
|  |  | Perceived healthcare benefit |  |  |

### Facilitators

The Andersen-Newman theoretical framework delineates facilitating factors as elements related to the logistical aspects of accessing healthcare [21]. These factors create conditions that empower an individual to align with a particular value or fulfill a health-related need, influencing their utilization of health services.

### Clinic environment

Participants in the study identified structural clinic environment facilitators as key contributors to the reliability of PHC services, serving as motivation for older individuals to seek and access care.

#### Reliable PHC services

Older people expressed that public hospitals were their preferred choice for accessing primary health care services due to the perceived reliability of these services. Some older individuals highlighted the consistent availability of medications for their multimorbidity as a compelling factor, serving as a significant motivation for them to consistently choose healthcare facilities for their care. Public hospitals were favored primarily due to their non-imposition of user fees. This preference stemmed from the fact that a significant majority of older people lacked a stable source of income to cover associated medical expenses:

> *“The quality of care here is good. We don’t pay for any services; we get the services for free.”* (Female, age range 76-80, patient, Mzimba).

Caregivers, often accompanying older individuals to public health facilities, shared similar sentiments. They utilized this experience as a tool to encourage the older people to consistently access (PHC) services.

> *“……on that one, it would encourage them because at this hospital, here at the central hospital (KCH), where we are, it is the most reliable hospital in this district and so we still try to come here to receive care”.* (Female, age range 36-40, caregiver, Lilongwe).

### Social factors

Various factors motivated older people to utilize PHC services. The availability of transportation support, combined with their perceived healthcare benefits, served as the primary drivers that encouraged them to continue accessing PHC.

#### Available means of transportation

The considerable distance to PHC facilities posed a significant challenge for older individuals, making it difficult for them to either walk (due to physical limitations) or afford transportation (due to an unreliable income source). Consequently, any external support, whether in the form of cash or kind, for transportation was greatly appreciated:

> *“What I know is that the hospital collaborates with MSF (Médecins Sans Frontières) through a project which economically empowers the patients especially those with cancer of the cervix in terms of transportation to and from the hospital”.* (Palliative care clinic nurse, Mangochi)

#### Perceived health care benefit

Older people derived extra motivation, either from personal satisfaction stemming from perceived healthcare benefits or through encouragement from both family members and healthcare workers:

> *“No, the hospital is my refuge. It’s my source of assistance, and what they say is accurate. The agony I endured from ulcers, had I opted for traditional medicine, I might not be alive today”.* (Female, age range 56-60, patient, Lilongwe).

## Barriers

While not prominently featured as a primary domain, barriers are acknowledged to coexist with the predisposing factors within the Andersen-Newman theoretical framework [23]. These factors serve as obstacles that hinder the access and utilization of healthcare services.

### Clinic environment

The structural clinic environment barrier, as perceived by participants, primarily revolves around the setup of PHC clinics.

#### Inconvenient clinic setup

Older individuals spent a significant amount of time at the clinics, leading to the need for restroom facilities that presented various challenges. Conversely, older people noted lack of a designated PHC operating space at Mangochi District Hospital. Consequently, they expressed experiencing frequent relocation within the facility, with some of these alternative locations deemed suboptimal for efficient service delivery.

> *“The lack of a toilet at our primary care facility has emerged as a significant issue for us”.* (Female, age range 51-55, patient, Mangochi).

### Geographical factors

The accessibility of PHC facilities at secondary or tertiary levels faced challenges, primarily attributed to geographical factors of either bad road conditions or the unavailability of primary care services at the community health centers.

#### Bad road conditions

Respondents expressed concern with the poor conditions of the roads, compounded by the considerable distance between their communities and the primary care facilities. This resulted in a significantly lengthy travel time, which was not conducive for the older people dealing with the consequences of their multimorbidity throughout the entire journey:

> *“The distance that we cover to access the primary health care is too long, the road that we use is not good either”.* (Male, age range 21-25, caregiver, Mangochi).

#### Lack of comprehensive PHC at the local health facility

Older people reported a deficiency in PHC services at community health centers, citing either a lack of skilled healthcare professionals or essential medicines and supplies to address their medical conditions. Consequently, they expressed the necessity to travel extensive distances to access the services at referral hospitals:

> *“The sad thing is that from the areas where we come from, clinics are there but when we go there, they always say that they don’t have medicine and test kits for the conditions that we have”.* (Female, age range 51-55, patient, Mangochi).

### Social factors

The interviews revealed a pattern among older individuals who turn to alternative medicine, driven by treatment fatigue at PHC facilities and a belief in its potential to provide healing for their various health conditions.

#### Use of alternative medicine

Various factors were identified as reasons why certain older individuals avoid utilizing PHC services. Among these factors is a preference for relying on herbal medicine and belief in faith healing:

> *“There are many herbal medicines nowadays and other things and so they are told to use such in the hopes that they would get better.”* (NCD clinic nurse, Lilongwe).

## Need factors

Need factors within the Andersen-Newman theoretical framework play a crucial role by representing solutions that service users believe can enhance service utilization.

### Clinic environment

The older people underscored the significance of dedicating resources to research initiatives. Their belief rested on the premise that research endeavors would uncover innovative approaches, ultimately leading to enhancements in their overall quality of life.

#### Research on diabetic foods

Older people and their caregivers strongly advocated for increased research on diabetic foods, with the expectation that a diverse range of affordable options would become available. This sentiment stemmed from the recognition of the economic constraints experienced by them. A significant number of older individuals lacked a stable source of income and depended on insufficient handouts, making it challenging to afford the diabetic foods available on the market. Consequently, adhering to the recommended diabetic diet seemed like an unattainable goal for them:

> *“The researcher should also do research into the types of food that people with diabetes can afford so that we do not end up malnourished”.* (Male, age range 56-60, patient, Lilongwe).

### Geographical factors

The absence of PHC services at local health centers posed significant challenges for older individuals. Their physical conditions often made it impractical for them to walk to secondary or tertiary hospitals to access PHC. Additionally, the financial strain associated with paying for transportation presented a persistent obstacle. In response to these challenges, older people recommended the implementation of community-based approaches as a sustainable solution to address this bottleneck.

#### Community approaches

Respondents proposed either capacitating local health centers with expert personnel and resources to manage primary care conditions at local level or having scheduled community visits by the hospital primary care teams to attend to patients at the community health facilities:

> *“I believe that receiving care and medications at our local health centers would be a highly commendable intervention if it were feasible”.* (Female, age range 56-60, caregiver, Mzimba).

### Social factors

To foster self-reliance and improve their quality of life, older individuals voiced a robust desire for assistance in the form of startup capital for small-scale business ventures.

#### Business support

Acknowledging the ongoing economic challenges, older individuals and their caregivers proposed their inclusion in cash transfer initiatives or similar loan facilities offered by various institutions to initiate small-scale business ventures. They expressed optimism that such interventions could establish a reliable source of income, consequently ensuring food security at home and alleviating transportation and other related challenges when accessing PHC services at various health facilities:

> *“Older people should also get support to have startup capital to run small-scale businesses in order to be self-reliant but also facilitate ease of access and utilization of primary health care services”.* (Female, age 55-60, patient, Mangochi).

## Discussion

The Andersen-Newman theoretical framework of health services utilization was useful in illuminating several factors influencing accessibility of PHC services by older people with progressive multimorbidity in Malawi [21]. Older people may or may not access PHC for a range of reasons, spanning from personal factors to issues within the healthcare system [7].

Our research demonstrated that access to and utilization of primary healthcare services by older individuals are influenced by a combination of barriers, facilitators, and need factors. These factors can be classified into three overarching thematic categories: clinic environment, geographical factors and social factors. In general, the barriers, facilitators, and need factors exhibited substantial similarity across the three study sites. This likeness can be attributed to the public nature of these facilities, where services were offered free of charge at the point of delivery [23,24]. Consequently, the shared aspect of providing services without cost likely contributes to comparable experiences across the sites.

Barriers identified in this research align with the results of a systematic review of prior studies that have identified barriers impacting the accessibility and utilization of primary healthcare services among older people living with progressive multimorbidity in LMICs [9]. In all three study locations, a recurring observation was the absence of comprehensive PHC services at the community health facilities. This aligns with the results of a previous study conducted in Nigeria [25]. These facilities primarily served as referral points to the next level as they lacked both expert PHC providers and quality essential medicines and supplies [26]. A referral to the higher-level facility (secondary or tertiary) was also met with additional geographical barriers that arose due to poor road conditions. This not only made travel uncomfortable for older individuals but also prolonged the journey significantly. The referral hospital’s inconvenient clinic setup, marked by poorly designed infrastructure, presented an extra barrier for the older people as described elsewhere [3,27,28]. In contrast to the two other study sites, older people in Mangochi faced challenges in adapting to the ongoing makeshift administrative arrangements concerning the venues for PHC clinics at the hospital. Due to enduring such exasperating administrative ordeals, some older people turned to alternative medicine, opting for either traditional and herbal remedies or seeking solace in faith healing practices. The research substantiates the overarching idea that numerous contextual barriers hinder the access and utilization of primary healthcare services by older people [9].

The identified facilitating factors align with prior research in resource-limited settings, reflecting a concurrent trend [9]. Nevertheless, our study has unveiled novel insights. Specifically, we found that the perceived benefits of healthcare played a crucial moderating role in shaping individuals’ navigation through various barriers. Notably, satisfaction with advancements in personal health outcomes emerged as a significant motivating factor, driving the ongoing utilization of PHC [29]. The economic hurdles linked to transportation and associated healthcare costs disproportionately affected older individuals, imposing a substantial burden [10,11,30]. Recognizing this issue, certain partner organizations, such as Médecins Sans Frontières (MSF), collaborated with the Ministry of Health to alleviate the challenges faced by older people in Mangochi District. They extended assistance by offering transport support for scheduled monthly clinic visits, thereby enhancing access to healthcare [31]. This support underscores Mangochi District’s capacity to meet a significant demand for PHC for older people with cervical cancer, with a notably higher proportion compared to Mzimba and Lilongwe, where a comparable level of support appears to be absent. Similarly, all the study sites, being public health facilities, did not charge user fees. This healthcare financing mechanism remains applicable to most LMICs following revelations that user fees serve as a barrier to accessing health services [32]. Moreover, our study findings corroborate this, underscoring it as a notable determinant of older peoples’ access to PHC.

Strikingly, as a crucial requirement, older people expressed the need for assistance in acquiring startup capital for establishing small-scale businesses. This initiative aims to enhance financial independence, consequently facilitating access to and utilization of PHC services. As such, they advocated for their inclusion in social cash transfer initiatives, typically overlooked, or sought opportunities to access loan facilities. This proposition aligns with the World Health Organization’s (WHO) report on aging and health, which advocates for the implementation of social assistance measures, including social pensions and related social protection initiatives, in favor of older people [33]. Emphasis should however be placed on the importance of cultivating the capabilities of the older people by allocating resources to their healthcare and long-term support. The objective is to optimize their functional capacity, sustain productive work, and enhance overall health-related quality of life [34].

These findings should be considered within the broader context of general deficiencies observed in PHC settings in Malawi. PHC facilities seemed to be entrenched in inflexible structures, where the understanding, or lack thereof, regarding diverse challenges faced by older people or service delivery had minimal impact on driving enhancements in service delivery. Paradoxically, this is happening despite the Malawian government’s active promotion of the implementation of the National Policy for Older Persons. [7]. The policy aims to improve elderly-friendly health services and advocates for its seamless integration into existing health facilities, with the goal of enhancing the management of chronic diseases associated with aging. This is in accordance with the World Health Organization’s emphasis on healthcare centered around individuals [35]. It involves incorporating their perspectives and acknowledging them as both participants and beneficiaries within dependable health systems. In the midst of an economic downturn, it is crucial for the Malawi government to foster increased collaboration with relevant stakeholders. This is necessary to build a robust PHC foundation that can effectively address the diverse care needs of older people living with progressive multimorbidity [12].

The results of this research offer supplementary insights into the state of PHC for older people with multimorbidity in Malawi, carrying implications for policies and practices in LMICs.

### Strengths and limitations

The present study exhibits various strengths, particularly its multi-site approach and the involvement of diverse stakeholders across study locations. This methodology bolsters the study’s robustness by providing a comprehensive contextual understanding of the reported experiences of older individuals living with progressive multimorbidity. However, some older people and their caregivers might have viewed research assistants as integral members of the primary care team. This potential perception could introduce a power imbalance, leading participants to respond differently than they typically would, thereby posing a threat to the internal validity of the data. Nevertheless, the overall consistency in responses helps alleviate this concern.

We suggest that forthcoming studies involve hospital management teams, Ministry of Health officials responsible for clinical, nursing, and public health services, and politicians, as they play a central role in the policy formulation processes. Nonetheless, we assert the validity of the findings presented in this paper. This confidence is grounded in the triangulation of our results, achieved through interviews with diverse categories of stakeholders across various regions and districts of Malawi, encompassing urban, peri-urban, and rural communities.

## Supporting information

S1 Fig 1. The Andersen Newman Theoretical framework of health services utilization.

(DOCX)

S2 File. Distress Protocol.

(XLSX)

## Author Contributions

**Conceptualization**: Duncan Kwaitana, Eric Umar.

**Data curation:** Duncan Kwaitana, Eric Umar, Esnath Msowoya.

**Formal analysis:** Duncan Kwaitana, Eric Umar, Esnath Msowoya.

**Funding acquisition:** Thomas Mildestvedt, Eivind Meland, Maya Jane Bates.

**Investigation:** Duncan Kwaitana, Maya Jane Bates, Dorothee van Breevoort, Thomas Mildestvedt, Eivind Meland.

**Methodology:** Duncan Kwaitana, Eric Umar, Maya Jane Bates, Dorothee van Breevoort, Thomas Mildestvedt, Eivind Meland.

**Project administration:** Duncan Kwaitana, Eric Umar.

**Supervision:** Eric Umar, Maya Jane Bates, Dorothee van Breevoort, Thomas Mildestvedt, Eivind Meland.

**Validation:** Duncan Kwaitana, Eric Umar.

**Visualization:** Duncan Kwaitana, Eric Umar

**Writing – original draft:** Duncan Kwaitana, Eric Umar.

**Writing – review & editing:** Duncan Kwaitana, Eric Umar, Maya Jane Bates, Dorothee van Breevoort, Thomas Mildestvedt, Eivind Meland.

## Funding

This research received financial backing from NORHED PRICE, administered by the University of Bergen in Norway, and the Royal Society of Tropical Medicine and Hygiene. Funders played no part in shaping the design or content of this manuscript.

## Competing interests

The authors have declared that no competing interests exist.

## Data Availability

The qualitative data gathered for this research has been included as supplementary information. Please find it in S1 File, which contains subthemes derived from the qualitative data along with their respective definitions.

---

**Fig 1:**
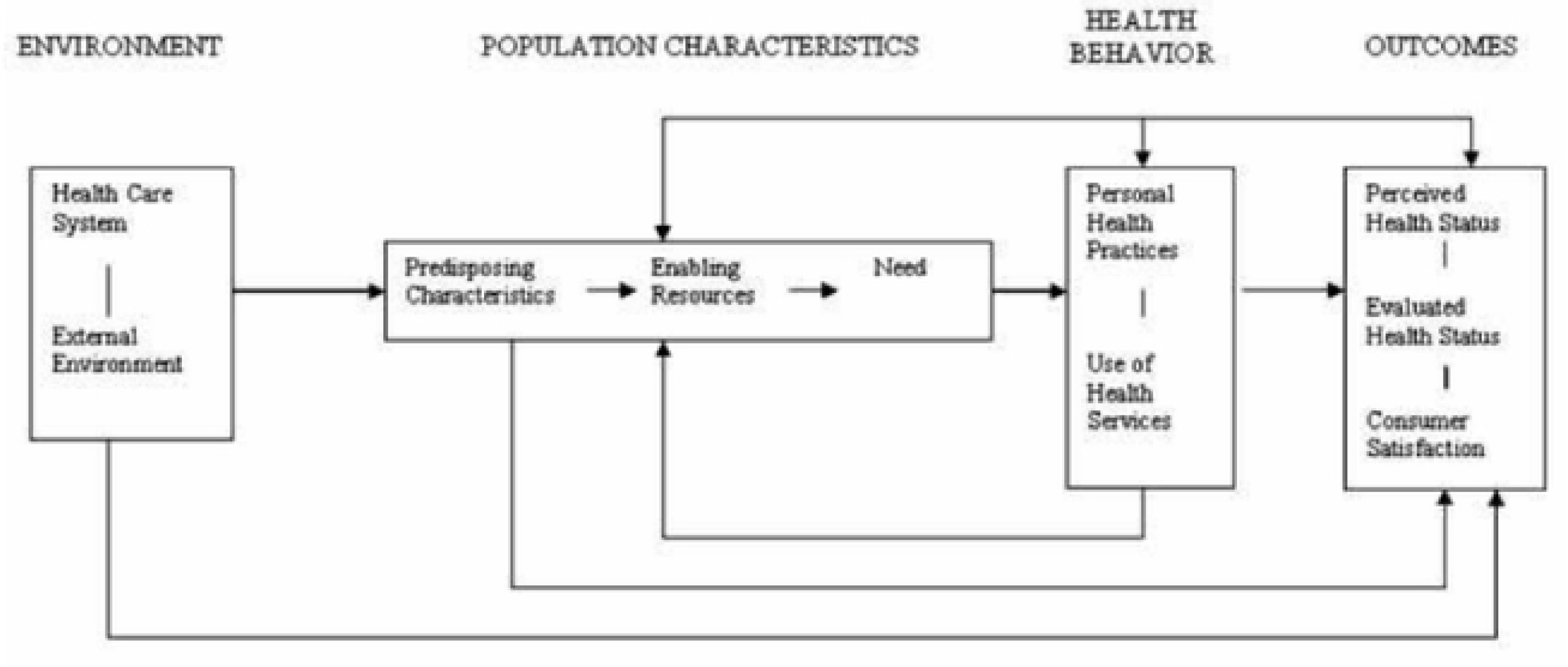
The Andersen-Newman Theoretical Framework of HealthServices Utilizations.

## Notes

### Competing Interest Statement

The authors have declared no competing interest.

### Author Declarations

Ethical approval for the study was granted by the College of Medicine Research and Ethics Committee (COMREC) with the assigned application number P.05/22/3646.

